# Efficacy and Safety of Pemvidutide in Patients with Metabolic Dysfunction-Associated Steatohepatitis: A Systematic Review and Dose-Specific Meta-Analysis of Randomized Controlled Trials

**DOI:** 10.64898/2025.12.22.25342824

**Authors:** Ehab Abdu, Raghad Alghamdi, Alaa Mohsen, Zahra Alabbad, Mazin Kharabah, Anas Al-Thagfi, Mawadah Al-Shabeeb, Faisal Alanzi, Hanan Al-Harthi, Ali Alessa, Ahmed Al-Murdhimah

## Abstract

**Background:** Metabolic dysfunction–associated steatohepatitis (MASH) is a prevalent and progressive liver disease with limited pharmacologic treatment options. Pemvidutide, a GLP-1–glucagon dual receptor agonist, has shown promise in targeting both hepatic steatosis and fibrosis. This systematic review and meta-analysis aimed to evaluate the efficacy and safety of pemvidutide in adults with MASH.

**Methods:** We systematically searched PubMed, Scopus, Web of Science, and Cochrane CENTRAL through December 15, 2025, for randomized controlled trials (RCTs) comparing once-weekly subcutaneous pemvidutide (1.2 mg, 1.8 mg, or 2.4 mg) with placebo in adult MASH patients. Primary outcomes were changes in liver fat content (LFC, % via MRI-PDFF) and Enhanced Liver Fibrosis (ELF) score. Secondary outcomes included body weight, glycemic and lipid parameters, blood pressure, heart rate, and adverse gastrointestinal events. Dose-specific pairwise meta-analyses were performed using a random-effects model.

**Results:** Three RCTs encompassing 370 participants (195 pemvidutide, 175 placebo) were included. Pemvidutide significantly reduced LFC at all doses, with the greatest effect at 1.8 mg (MD = −21.63%, 95% CI: −27.23 to −16.02; p<0.0001). ELF score improvement was significant at 1.2 mg and 1.8 mg doses but not at 2.4 mg. Across all doses, body weight decreased significantly, while HbA1c remained unchanged. Pemvidutide also reduced systolic blood pressure and total cholesterol (1.8 mg and 2.4 mg), with modest HDL reduction at 2.4 mg. Mild-to-moderate gastrointestinal adverse events were observed, with nausea more frequent at 1.8 mg.

**Conclusions:** Pemvidutide is effective in reducing liver fat and improving cardiometabolic parameters in MASH, with a favorable safety profile. Dose-specific effects on fibrosis suggest potential early antifibrotic activity, highlighting its promise as a dual-targeted therapy for MASH. Further long-term studies are warranted to confirm sustained hepatic and metabolic benefits.

## 1. Introduction

Metabolic dysfunction–associated steatohepatitis (MASH) represents a substantial global health burden, characterized by ≥ 5% hepatic steatosis, affecting up to 38% of the global population and 80% of individuals with obesity [1–4]. MASH is the hepatic manifestation of metabolic syndrome and is strongly associated with obesity, insulin resistance, and type 2 diabetes mellitus, with significant implications for cardiovascular, oncologic, and liver-related morbidity and mortality [5]. Until 2024, no pharmacologic therapies were approved for the treatment of MASH. Resmetirom, a thyroid hormone receptor-β agonist and the first therapy approved by the U.S. Food and Drug Administration (FDA) for MASH, has demonstrated efficacy in resolving steatohepatitis without worsening fibrosis and in improving fibrosis without exacerbating MASH. However, it does not produce clinically significant weight loss beyond that attainable with lifestyle modification, including diet and exercise [6]. Glucagon-like peptide-1 receptor (GLP-1R) agonist monotherapy has also been shown to induce resolution of MASH, but it has not demonstrated statistically significant improvements in hepatic fibrosis, as its benefits are primarily mediated indirectly through reductions in liver fat content (LFC) driven by weight loss [7,8]. Although both approved monotherapies are effective in the treatment of MASH resolution, a combined therapeutic approach may provide both rapid improvements in MASH and greater reductions in body weight. Preliminary trials suggested the potential benefit of pemvidutide, a GLP-1–glucagon dual receptor agonist, in MASH resolution and fibrosis improvement, by targeting MASH pathophysiology through glucagon-mediated hepatic effects, weight reduction, and anti-inflammatory actions via GLP-1 [9].

Therefore, high-quality pooled evidence is needed to confirm the efficacy and safety of pemvidutide and to support robust clinical recommendations; in this systematic review and meta-analysis, we aimed to evaluate its therapeutic impact in patients with MASH.

## 2. Method

This systematic review and meta-analysis was conducted in accordance with the Preferred Reporting Items for Systematic Reviews and Meta-Analyses (PRISMA) guidelines [10]. All methodological procedures adhered to the recommendations outlined in the Cochrane Handbook for Systematic Reviews of Interventions [11]. The study protocol was prospectively registered in the International Prospective Register of Systematic Reviews (PROSPERO 2025; registration number: CRD420251270536).

### 2.1 Search Strategy and Study Selection

We conducted a comprehensive and systematic search of major electronic databases, including PubMed, Scopus, Web of Science, and Cochrane Central Register of Controlled Trials (CENTRAL), from inception to December 15, 2025. Eligible studies had to compare once-weekly subcutaneous pemvidutide (1.2 mg, 1.8 mg, or 2.4 mg) with placebo in patients diagnosed with metabolic dysfunction–associated steatohepatitis (MASH). A highly sensitive search strategy was developed using Boolean operators and a combination of Medical Subject Headings (MeSH) and free-text terms. The core search syntax included: (“pemvidutide” OR “GLP-1 glucagon dual receptor agonist” OR “glucagon-like peptide-1 glucagon dual agonist“) AND (“Metabolic Dysfunction-Associated Steatohepatitis” OR “MASH” OR “Non-alcoholic Steatohepatitis” OR “NASH“). To ensure completeness, we also manually screened the reference lists of all eligible articles and relevant systematic reviews for additional studies. The full search strategies for each database are provided in **Table S1.** All retrieved records were imported into EndNote for duplicate removal. Following deduplication, the proposed studies were uploaded into the Rayyan screening platform for further evaluation [12]. Two independent reviewers screened titles and abstracts, followed by full-text assessment of potentially eligible studies based on predefined PICO criteria. Any discrepancies were resolved through discussion with a third reviewer.

### 2.2 Eligibility Criteria and Clinical Outcomes

We included RCTs with full-text articles published in English that evaluated the efficacy and safety of once-weekly subcutaneous pemvidutide, administered at any investigated dose (including 1.2 mg, 1.8 mg, or 2.4 mg, with or without dose titration), compared with placebo in adult patients (≥18 years) with metabolic dysfunction–associated steatohepatitis (MASH). Eligible studies enrolled participants with confirmed hepatic steatosis or steatohepatitis assessed by magnetic resonance imaging–proton density fat fraction (MRI-PDFF) and/or liver biopsy, with or without liver fibrosis. We excluded observational studies, non-randomized trials, animal or in-vitro studies, reviews, conference abstracts without full data, and studies enrolling patients with other chronic liver diseases, significant alcohol consumption, or decompensated cirrhosis. The primary efficacy outcomes were changes from baseline in liver fat content (%) assessed by magnetic resonance imaging–proton density fat fraction (MRI-PDFF) and change in the Enhanced Liver Fibrosis (ELF) score. Secondary outcomes included changes in body weight; glycemic parameters, specifically glycated hemoglobin (HbA1c); lipid profile parameters, including total cholesterol, low-density lipoprotein cholesterol (LDL), high-density lipoprotein cholesterol (HDL), and triglycerides; and hemodynamic parameters, including systolic blood pressure (SBP), diastolic blood pressure (DBP), and heart rate (HR). Safety outcomes comprised gastrointestinal adverse events, categorized as mild or moderate, including nausea, diarrhea, and constipation. All outcomes were evaluated using parallel, dose-specific pairwise meta-analyses comparing pemvidutide doses of 1.2 mg, 1.8 mg, and 2.4 mg with placebo.

### 2.3 Data Extraction and Risk of Bias Assessment

Data extraction was performed independently by two reviewers using a standardized data collection form developed in Microsoft Excel. Extracted information included study characteristics, patient demographics, and reported outcomes. To ensure accuracy, all extracted data were cross-checked for discrepancies. The risk of bias of included RCTs was assessed using the Cochrane Risk of Bias 2 (RoB 2) tool [13], evaluating bias arising from the randomization process, deviations from intended interventions, missing outcome data, measurement of outcomes, selection of the reported results, and other potential sources of bias. Each study was classified as having a low risk of bias, some concerns, or a high risk of bias. Any disagreements between reviewers were resolved through discussion with a third reviewer.

### 2.4 Statistical Analysis and Heterogeneity Assessment

All statistical analyses were performed using R software (version 4.5.1). For continuous outcomes, pooled treatment effects were estimated as mean differences (MDs) with corresponding 95% confidence intervals (CIs). Meta-analyses were conducted using a random-effects model based on the DerSimonian–Laird method to account for anticipated clinical and methodological heterogeneity among the included studies. Parallel pairwise meta-analyses were performed, and prespecified subgroup analyses were conducted according to intervention dose. Statistical heterogeneity was assessed using Cochran’s Q test and quantified using the I² statistic, with values of approximately 25%, 50%, and 75% representing low, moderate, and high heterogeneity, respectively. Between-study variance was estimated using the τ² statistic. Differences between subgroups were evaluated using the χ² test for subgroup differences. All statistical tests were two-sided, and a p-value < 0.05 was considered statistically significant. Meta-analysis results were presented graphically using forest plots.

## 3. Results

### 3.1 Literature Search Results

The systematic literature search across four databases (PubMed, Scopus, Cochrane, and Web of Science) identified a total of 186 records. After removing 9 duplicates, 177 unique records underwent title and abstract screening. Of these, 154 were excluded, leaving 23 full-text articles for eligibility assessment based on predefined criteria. Following detailed evaluation, 20 articles were excluded for the following reasons: 6 were conference reports, 3 were review articles, 2 were notes or extensions of existing studies, and 9 did not meet the inclusion criteria. Ultimately, 3 studies were deemed eligible and included in the final systematic review and meta-analysis [14–16]. The detailed selection process is illustrated in **Figure 1**.

**Figure 1.**
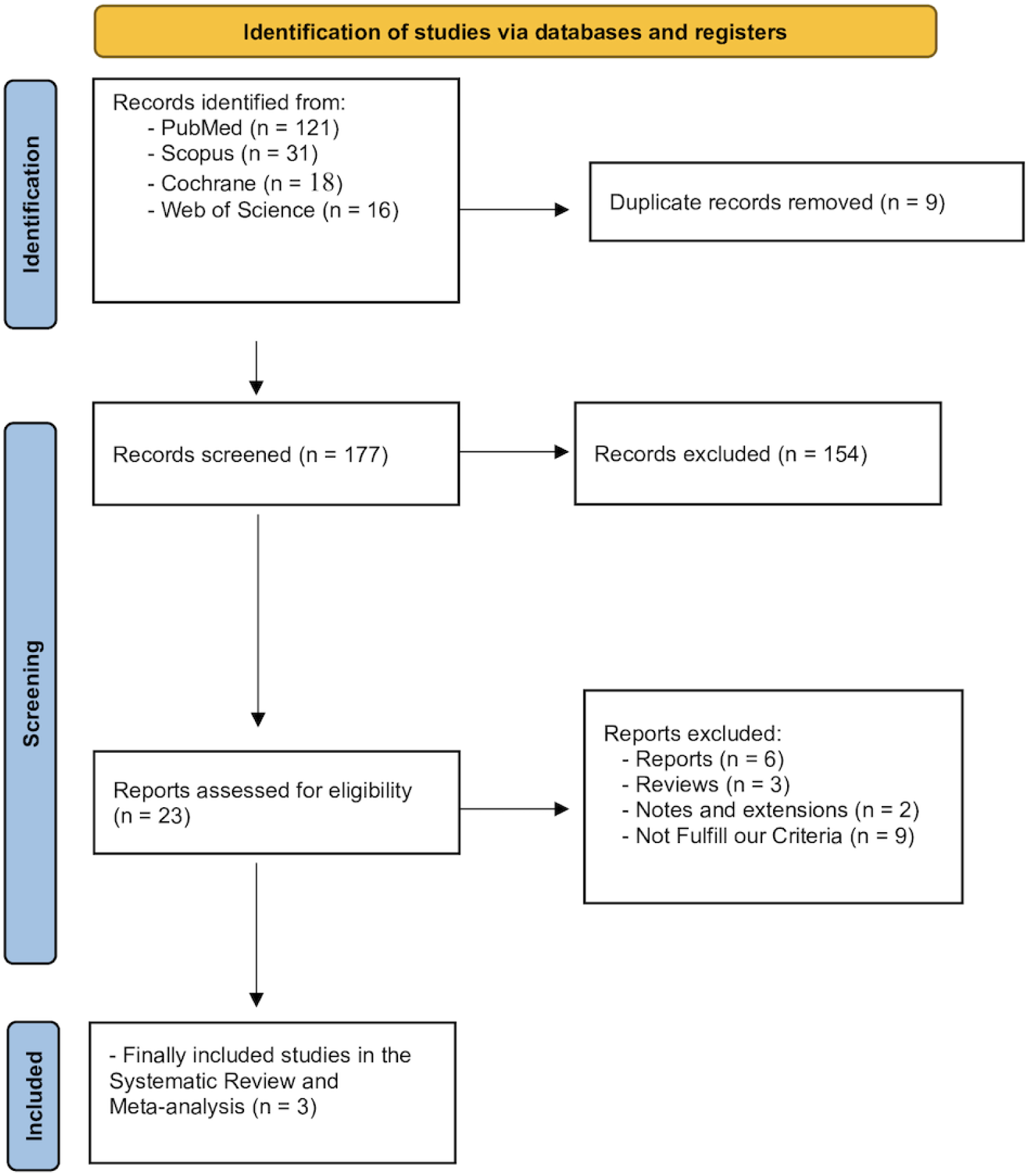
PRISMA flow diagram

### 3.2 Characteristics of Included Studies and Risk of Bias Assessment

This meta-analysis included RCTs published between 2024 and 2025, with a total pooled sample of 370 participants (195 in the pemvidutide group and 175 in the placebo group). The studies were primarily conducted in the USA, with one trial (IMPACT 2025) [14] also including sites in Australia. All trials assessed the efficacy and safety of pemvidutide administered as a once-weekly subcutaneous injection across various dose regimens (1.2 mg, 1.8 mg, and/or 2.4 mg) compared to placebo, with follow-up durations of 12 or 24 weeks. Participants were adults aged 18–75 years with obesity (BMI ≥27–28 kg/m²) and significant liver disease. Detailed baseline characteristics and summaries of the studies included are presented in **Tables 1 and 2**. The risk of bias assessment using the ROB-2 tool indicated that two studies had an overall low risk of bias, whereas the Harrison 2024 study [15] The study raised some concerns **(Figure 2).**

**Figure 2.**
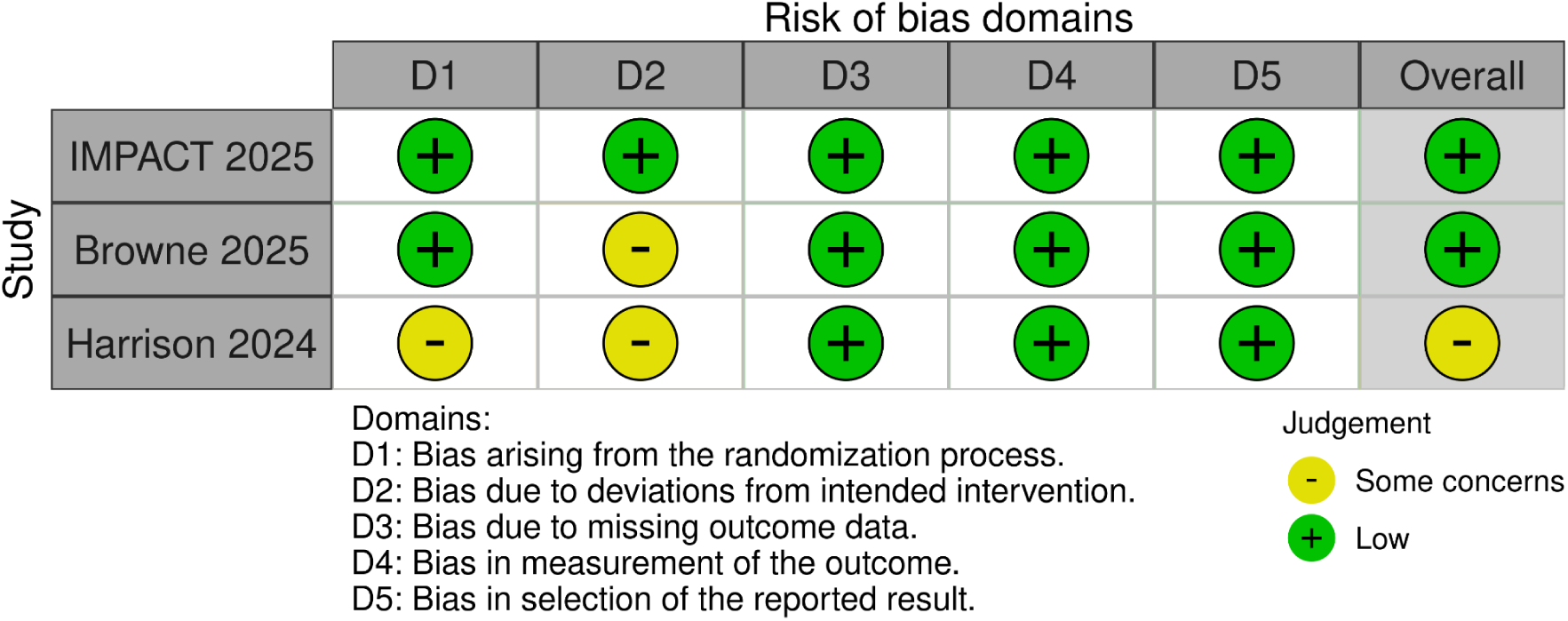
Risk of bias assessment (ROB-2).

**Table 1:** Summary Characteristics of included studies.

| Study ID | Arms | Sample Size | Study design | Country | doses (mg, weekly) | Follow up | Conclusions |
| --- | --- | --- | --- | --- | --- | --- | --- |
| IMPACT 2025 | Pemvidutide | 126 | RCTs | USA and Australia | 1.2 mg or 1.8 mg administered subcutaneously once weekly | 24 w | Pemvidutide treatment met the primary endpoint of MASH resolution without worsening of fibrosis at 24 weeks. It did not meet the primary endpoint of fibrosis improvement without worsening of MASH at this timepoint. The drug was well tolerated, with low discontinuation rates, despite the absence of dose titration. |
|  | Placebo | 86 |  |  |  |  |  |
| Browne 2025 | Pemvidutide | 45 | RCTs | USA | 1.2 mg, 1.8 mg, or 2.4 mg administered subcutaneously once weekly | 24 w | 24 weeks of pemvidutide treatment resulted in significant reductions in liver fat content (up to 76.4% relative reduction) and body weight (up to 6.2% loss) that further improved upon effects observed at 12 weeks. The drug was well-tolerated with low incidence of side effects. |
|  | Placebo | 19 |  |  |  |  |  |
| Harrison 2024 | Pemvidutide | 24 | RCTs | USA | 1.2 mg, 1.8 mg, or 2.4 mg administered subcutaneously once weekly | 12 w | Weekly pemvidutide treatment yielded significant reductions in liver fat content (up to 68.5% relative reduction), markers of hepatic inflammation (ALT), and body weight compared to placebo. |
|  | Placebo | 70 |  |  |  |  |  |

**Table 2:** Baseline Characteristics of included studies.

| Study ID | Arms | Sample Size | Age (y), mean (SD) | Male, n (%) | Body weight (kg), mean (SD) | body mass index (kg/m<sup>2</sup>), mean (SD) | liver fat content (%), mean (SD) | ALT (IU/L), mean (SD) | AST (IU/L), mean (SD) | HbA1c (%), mean (SD) | DM, n (%) |
| --- | --- | --- | --- | --- | --- | --- | --- | --- | --- | --- | --- |
| **IMPACT 2025** | Pemvidutide 1.2 mg | 41 | 55.2 (13) | 16 (39) | 110.7 (26.3) | 39.2 (8.4) | 20 (7.1) | 67.6 (54.6) | 50.8 (26.8) | 6.3 (1.1) | 19 (46) |
|  | Pemvidutide 1.8 mg | 85 | 53.4 (12.4) | 35 (41) | 107.7 (21.3) | 38.7 (6.9) | 19 (6.8) | 67.6 (43) | 51 (29.4) | 6.5 (1.2) | 36 (42) |
|  | Placebo | 86 | 53.4 (12.4) | 38 (44) | 109.1 (25.1) | 38.7 (7.8) | 19.6 (6.4) | 56.6 (32.7) | 44.9 (20.8) | 6.4 (1) | 37 (43) |
| **Browne 2025** | Pemvidutide 1.2 mg | 16 | 48.6 (11) | 9 (56.3) | 100.9 (13.2) | 36 (3.8) | 20.1 (7.7) | 35.3 (13) | 23.6 (6.9) | 5.7 (0.3) | 3 (18.8) |
|  | Pemvidutide 1.8 mg | 15 | 49.9 (10) | 7 (46.7) | 101.4 (16.3) | 36.7 (6.1) | 23.9 (7.4) | 32.4 (14.2) | 24.4 (6.7) | 5.7 (0.3) | 6 (40) |
|  | Pemvidutide 2.4 mg | 14 | 48.4 (8) | 6 (42.9) | 107.4 (17.2) | 37.0 (5.3) | 20.5 (6.5) | 39.6 (26.6) | 29.4 (15.5) | 5.6 (0.4) | 3 (21.4) |
|  | Placebo | 19 | 49 (15) | 8 (42.1) | 104.4 (21.2) | 37.1 (4.9) | 24.0 (9.6) | 41 (21.3) | 25.1 (10.5) | 5.8 (0.2) | 5 (26.3) |
| **Harrison 2024** | Pemvidutide 1.2 mg | 23 | 48.6 (11) | 14 (60.9) | 102.4 (14.6) | 36.3 (5.6) | 21.6 (7.3) | 32.4 (13.8) | 25.4 (7.5) | 5.7 (0.3) | 7 (30.4) |
|  | Pemvidutide 1.8 mg | 23 | 50.3 (9) | 11 (47.8) | 98.9 (19.7) | 35.4 (3.9) | 21.8 (8) | 36.4 (15.6) | 24.6 (7.2) | 5.7 (0.3) | 7 (30.4) |
|  | Pemvidutide 2.4 mg | 24 | 48.8 (8) | 9 (37.5) | 98.2 (18.9) | 35.3 (5) | 20.2 (7) | 37.8 (24.4) | 27.2 (13.9) | 5.6 (0.4) | 7 (29.2) |
|  | Placebo | 24 | 47.9 (14) | 10 (41.7) | 105.1 (20.8) | 36.9 (4.7) | 23.8 (9.2) | 39.5 (21.4) | 23.8 (10) | 5.8 (0.2) | 6 (25) |

### 3.3 Primary Efficacy Outcomes: Parallel Pairwise Meta-Analysis

#### 3.3.1 Change in Liver Fat Content (%)

Dose-specific analysis demonstrated that Pemvidutide was associated with significant reductions in liver fat content compared to placebo across all evaluated doses. The greatest decrease was observed with the 1.8 mg dose (MD = −21.63, 95% CI: −27.23 to −16.02, p<0.0001; I^2^=96.4%, p<0.0001), followed by the 1.2 mg dose (MD = −14.17, 95% CI: −19.25 to −9.08, p<0.0001; I^2^=94.8%, p<0.0001), and 2.4 mg dose (MD = −12.47, 95% CI: −15.31 to −9.63, p<0.0001; I^2^=87.7%, p=0.004) **(Figure 3).**

**Figure 3.**
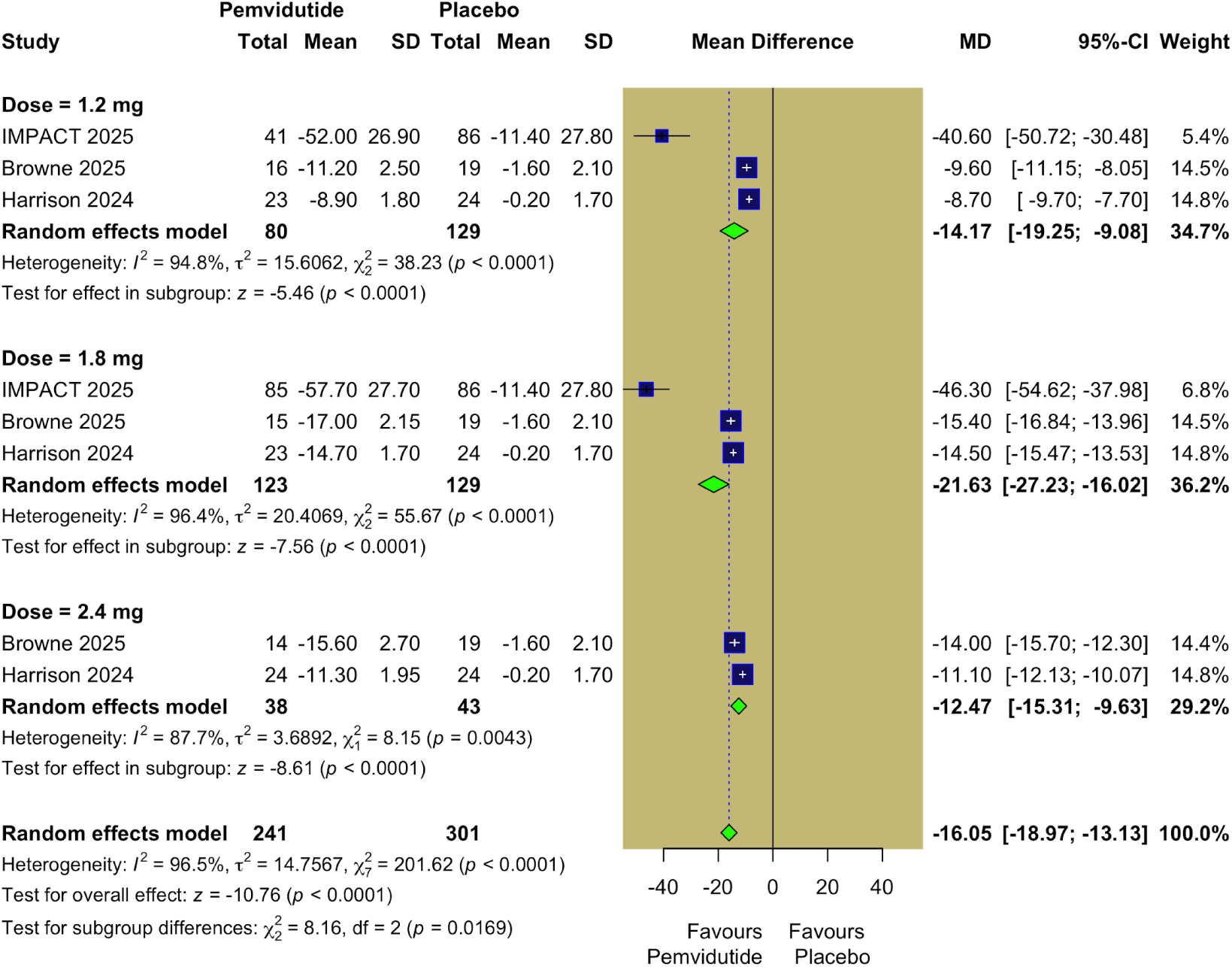
Change in Liver Fat Content (%).

#### 3.3.2 Change in Enhanced Liver Fibrosis (ELF) score

Dose-specific analysis demonstrated that Pemvidutide was associated with a significant reduction in the ELF score compared to placebo at the 1.2 mg dose (MD = −0.66, 95% CI: −1.17 to −0.14, p = 0.0122; I² = 88.7%, p = 0.0001) and the 1.8 mg dose (MD = −0.33, 95% CI: −0.58 to −0.09, p = 0.0076; I² = 51.1%, p = 0.1292), but not at the 2.4 mg dose (MD = −0.86, 95% CI: −2.43 to 0.70, p = 0.2792; I² = 94.9%, p < 0.0001) **(Figure 4).**

**Figure 4.**
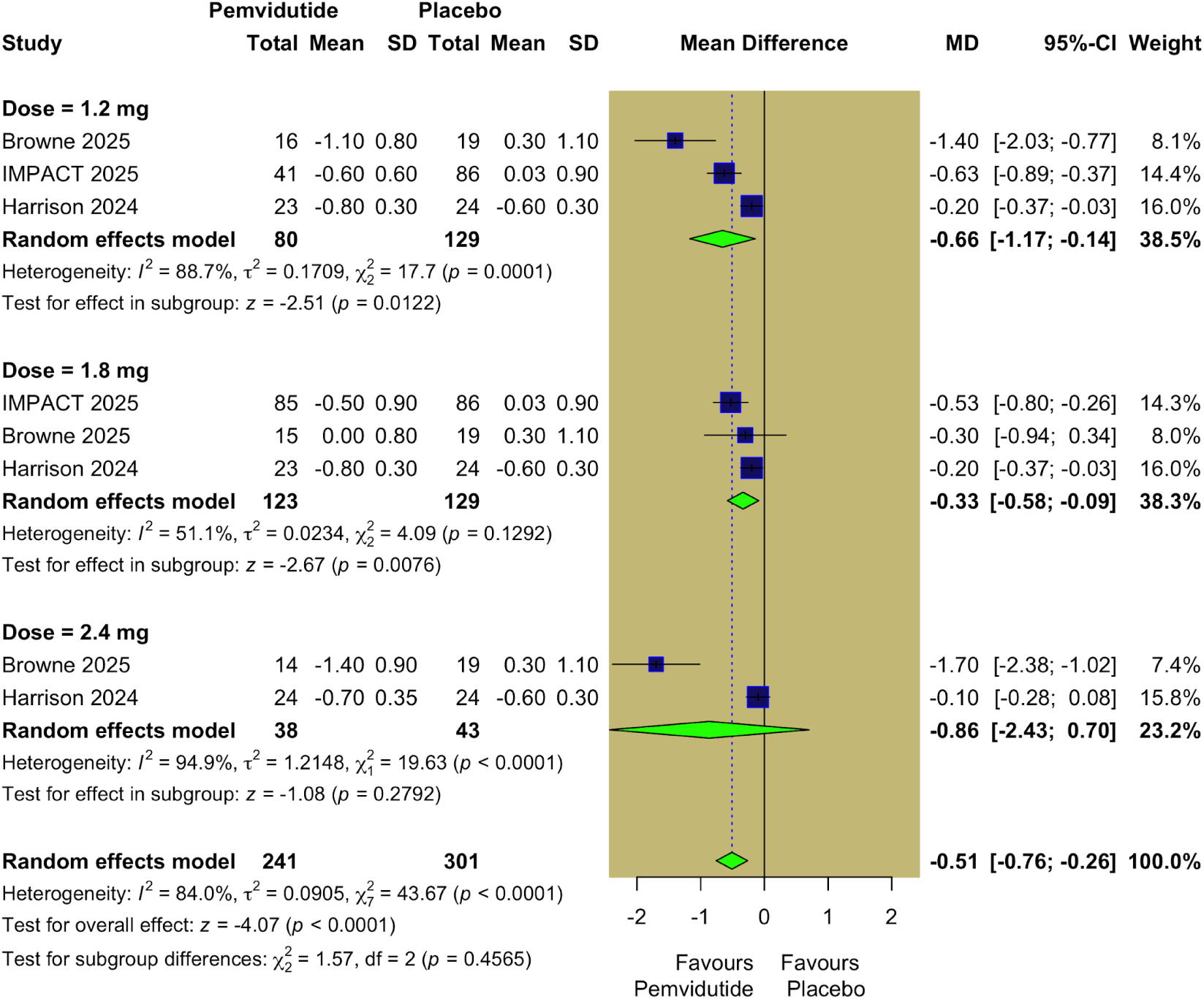
Change in Enhanced Liver Fibrosis (ELF) Score.

### 3.4 Secondary Outcomes: Parallel Pairwise Meta-Analysis

#### 3.4.1 Glycemic and Metabolic Outcomes

Dose-specific analysis demonstrated that Pemvidutide was associated with a significant reduction in body weight across all evaluated doses: 1.2 mg (MD = −3.56 kg, 95% CI: −4.14 to −2.98, p < 0.0001; I² = 49.8%, p = 0.1366), 1.8 mg (MD = −3.25 kg, 95% CI: −5.28 to −1.22, p = 0.0017; I² = 96.5%, p < 0.0001), and 2.4 mg (MD = −3.55 kg, 95% CI: −3.90 to −3.20, p < 0.0001; I² = 0%, p = 0.524) **(Figure 5).** In contrast, Pemvidutide did not demonstrate a statistically significant reduction in HbA1c compared to placebo across any of the evaluated doses (1.2 mg, 1.8 mg, and 2.4 mg) **(Figure S1).**

**Figure 5.**
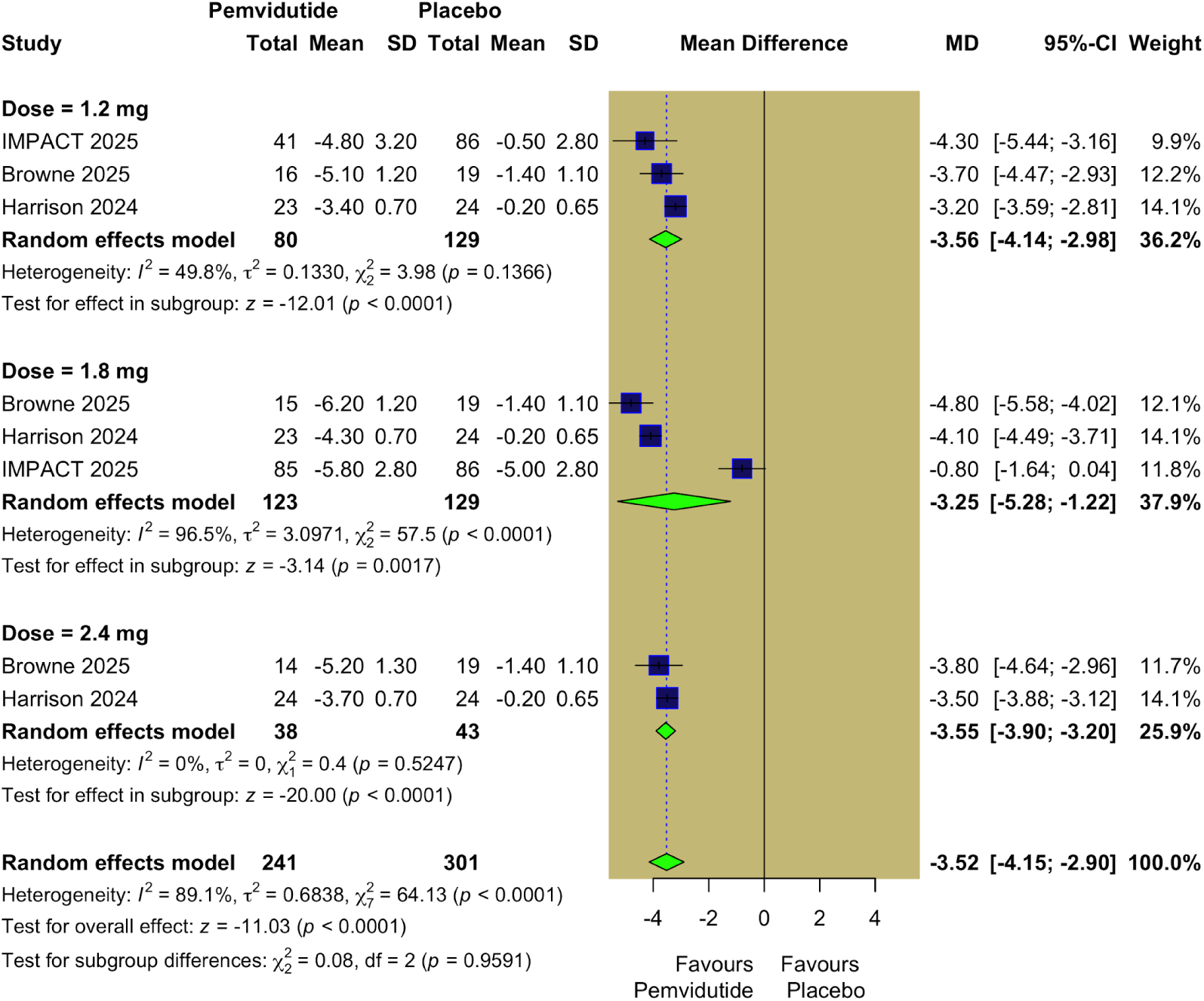
Change in Body Weight (kg).

#### 3.4.2 Atherogenic Lipid Parameters

Dose-specific analysis demonstrated that Pemvidutide was associated with a significant reduction in total cholesterol at the 1.8 mg dose (MD = −0.18 mmol/l, 95% CI: −0.28 to −0.08, p = 0.0004; I² = 0%, p = 0.5295) and the 2.4 mg dose (MD = −0.23 mmol/l, 95% CI: −0.35 to −0.11, p = 0.0001; I² = 10.6%, p = 0.2902), but was not significant at the 1.2 mg dose **(Figure S2).** For HDL cholesterol, a significant reduction was observed only at the 2.4 mg dose (MD = −0.03 mmol/l, 95% CI: −0.05 to −0.01, p = 0.0011; I² = 0%, p = 0.6616) but was not significant at the 1.2 mg and 1.8 mg doses **(Figure S3).** In contrast, pemvidutide did not demonstrate a statistically significant reduction effect on LDL cholesterol or triglycerides at any of the evaluated doses (1.2 mg, 1.8 mg, or 2.4 mg) **(Figure S4–S5).**

#### 3.4.3 Changes in Blood Pressure and Heart Rate

Dose-specific analysis demonstrated that Pemvidutide was associated with significant reductions in SBP at the 1.2 mg dose (MD = −7.98 mmHg, 95% CI: −9.09 to −6.87, p < 0.0001; I² = 0%, p = 0.3867), the 1.8 mg dose (MD = −4.88 mmHg, 95% CI: −7.11 to −2.65, p < 0.0001; I² = 69.4%, p = 0.0379), and the 2.4 mg dose (MD = −11.36 mmHg, 95% CI: −14.39 to −8.33, p < 0.0001; I² = 81.8%, p = 0.0192) **(Figure 6).** Moreover, Dose-specific analysis demonstrated that Pemvidutide was associated with a significant reduction in DBP was observed at the 1.8 mg dose (MD = −2.44 mmHg, 95% CI: −4.09 to −0.78, p = 0.0039; I² = 75.2%, p = 0.0178), whereas the 1.2 mg and 2.4 mg doses were not statistically significant **(Figure S6).** Additionally, pemvidutide was associated with a significant increase in the HR at the 1.2 mg dose (MD = 2.79 bpm, 95% CI: 0.62 to 4.96, p = 0.0117; I² = 84.1%, p = 0.0019), the 1.8 mg dose (MD = 1.13 bpm, 95% CI: 0.39 to 1.87, p = 0.0028; I² = 0%, p = 0.7548), and the 2.4 mg dose (MD = 1.57 bpm, 95% CI: 0.52 to 2.62, p = 0.0034; I² = 38.1%, p = 0.2036) **(Figure S7)**.

**Figure 6.**
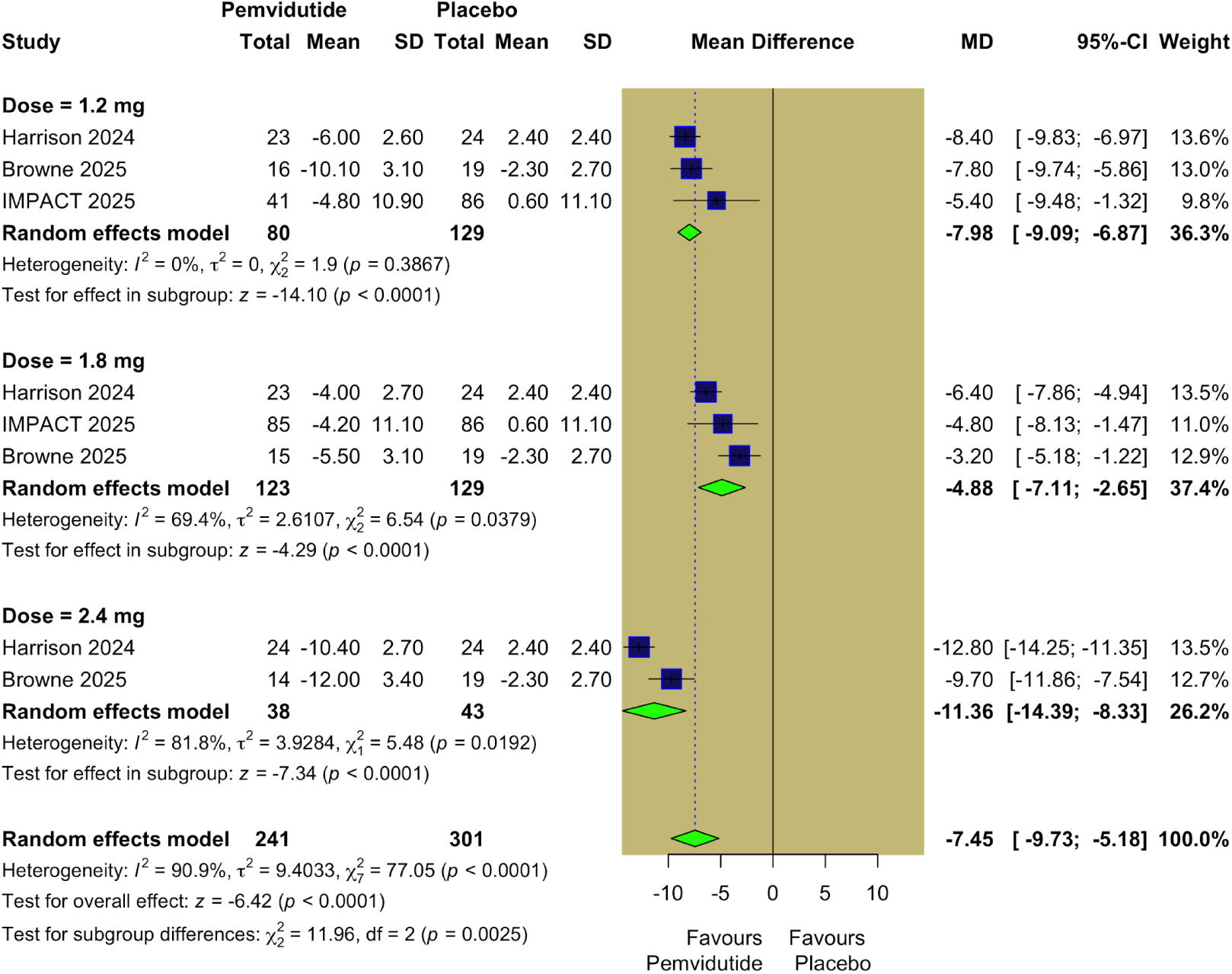
Change in Systolic Blood Pressure (mm Hg).

### 3.5 Gastrointestinal Adverse Events

#### 3.5.1 Mild adverse events

Dose-specific analysis indicated that Pemvidutide was not associated with a statistically significant increase in the incidence of diarrhea or constipation at any of the evaluated doses (1.2 mg, 1.8 mg, or 2.4 mg) **(Figure S8–S9).** In contrast, for nausea, a significant increase was observed at the 1.8 mg dose (RR = 2.92, 95% CI: 1.62 to 5.26, p = 0.0004; I² = 0%, p = 0.6685) but was not significant at the 1.2 mg and 2.4 mg doses **(Figure S10).**

#### 3.5.2 Moderate adverse events

Dose-specific analysis indicated that Pemvidutide was associated with a statistically significant increase in the incidence of diarrhea, constipation, or nausea at any of the evaluated doses (1.2 mg, 1.8 mg, or 2.4 mg) **(Figure S11–S13).**

## 4. Discussion

Our meta-analysis of three recent RCTs encompassing 370 patients represents the most comprehensive evaluation to date of pemvidutide, a GLP-1/glucagon dual receptor agonist for the treatment of MASH. In this dose-specific meta-analysis, pemvidutide demonstrated robust and clinically meaningful improvements in key hepatic, metabolic, and cardiometabolic parameters, supporting its therapeutic potential in patients with MASH. Pemvidutide was associated with significant reductions in liver fat content across all evaluated doses, with the greatest effect observed at the 1.8 mg dose. Improvements in the ELF score were evident at the 1.2 mg and 1.8 mg doses but not at 2.4 mg, indicating a dose-dependent but non-linear effect on fibrotic activity. Across all doses, pemvidutide produced consistent and clinically meaningful reductions in body weight; however, no statistically significant improvement in HbA1c was observed, suggesting that the observed hepatic and metabolic benefits were not primarily driven by glycemic control. With respect to lipid parameters, significant reductions in total cholesterol were observed at the 1.8 mg and 2.4 mg doses, while a modest reduction in HDL cholesterol was noted only at the highest dose, and no significant effects were detected on LDL cholesterol or triglycerides. Hemodynamically, pemvidutide significantly reduced systolic blood pressure across all doses, whereas diastolic blood pressure reduction was significant only at the 1.8 mg dose; a small but statistically significant increase in heart rate was observed across the dosing range. Overall, pemvidutide was generally well tolerated, with mild gastrointestinal adverse events comparable to placebo except for an increased incidence of nausea at the 1.8 mg dose, and no significant increase in moderate gastrointestinal adverse events at any dose.

The current Clinical Practice Guidelines for the diagnosis, treatment, and follow-up of individuals with MASH continue to focus on finding the optimal management for MASH. They recommend a multipronged approach encompassing lifestyle modifications, optimal comorbidity management, and targeted pharmacotherapy for specific high-risk groups [17]. Foundational to all treatment is lifestyle modification, which includes achieving sustained weight reduction to improve liver histology: >5% is needed to reduce liver fat, 7–10% to improve liver inflammation, and >10% to improve fibrosis, also limiting the consumption of ultra-processed food and sugar-sweetened beverages, and engaging in regular physical activity (preferably >150 min/week of moderate or 75 min/week of vigorous-intensity physical activity). Furthermore, alcohol consumption should be stopped completely and permanently in individuals with advanced fibrosis or cirrhosis [18]. For adults with non-cirrhotic MASH and significant liver fibrosis (stage ≥F2), a MASH-targeted treatment should be considered, if locally approved, with resmetirom being the recommended agent due to its demonstrated histological efficacy on steatohepatitis and fibrosis with an acceptable safety and tolerability profile in a large phase III trial [6,19]. For individuals with type 2 diabetes or obesity, optimal management of comorbidities is advised, often including the use of incretin-based therapies (such as semaglutide and tirzepatide), if indicated [20]. While GLP1 receptor agonists are safe to use in MASH (including compensated cirrhosis) when prescribed for their primary indications (T2D, obesity), they cannot currently be recommended as MASH-targeted therapies due to the absence of a formal histological improvement demonstration in large phase III trials [21].

MASLD is defined as the presence of excess triglyceride storage in the liver in the presence of at least one cardiometabolic risk factor, as well as fibrosis and cirrhosis [22]. Obesity plays an important role in the treatment of this condition as the main cause is excessive lipid storage in the entire body and especially in the liver, leading to histological features of hepatocellular ballooning and lobular inflammation. Thus, we pooled outcomes addressed these points. Our analysis demonstrates that pemvidutide significantly reduced LFC, liver fibrosis assessed by the ELF score, and promoted weight loss, and the cause of that is the dual receptor agonist on both GLP-1 and glucagon receptors; in contrast, a mono receptor agonist on GLP-1has demonstrated resolution of MASH, but it has not shown statistically significant improvements in fibrosis [7,23]. The cause of the superiority of pemvidutide to other GLP-1RAs on LFC and liver fibrosis is this effect on glucagon receptors, as its effect on the liver is stimulation of hepatic β-oxidation of fatty acids and a reduction of de novo lipogenesis. Additionally, it was hypothesized that due to coupled agonism, the same drug could achieve higher levels of liver fat reduction than those achievable by GLP-1R activation alone and have a greater effect on weight loss through reducing food consumption through anorectic effects and activation of glucagon receptors to increase energy expenditure, potentially giving similar effects to diet and exercise [24,25].

Our findings align with the first Phase 1b/2a trial (12 weeks) by Harrison 2025, which evaluated the effectiveness of pemvidutide for MASH patients; weekly subcutaneous doses of pemvidutide resulted in statistically significant reductions in hepatic fat compared to placebo, yielding up to a 68.5% relative reduction in LFC in only 12 weeks with The efficacy signal deepened in the 24-week Extension study, reaching a relative reduction of 75.2% [15]. Also showed a significant reduction in hepatic inflammation (serum ALT, cT1 relaxation time), and body weight. The Phase 2b IMPACT trial, while enrolling in a larger and more diverse population (n=212), corroborated these findings with a 57.7% relative reduction. The relative reduction in LFC was numerically superior in the smaller Extension trial (75.2%) compared to the larger IMPACT trial (57.7%)[14] .

In the phase 3 trial, the effectiveness of Semaglutide on MASH showed positive results on MASH resolution and fibrosis improvement, and although patients had significant weight loss, 72 weeks of treatment were required [26]. Additionally, a double-blind phase 2 trial involving patients with biopsy-confirmed MASH was conducted to evaluate the efficacy of semaglutide, which demonstrated the resolution of steatohepatitis but without fibrosis improvement [8]. It is possible that a combined therapeutic approach could provide both rapid effects on MASH and reductions in body weight.

Our analysis emphasizes a reduction in both systolic and diastolic blood pressure, thus giving a big benefit for MASH patients, as it leads to a decrease in cardiometabolic risk factors. The mechanism behind that is that weight loss plays a central role by reducing visceral adiposity, improving arterial compliance, and decreasing insulin resistance, all of which contribute to lower systemic vascular resistance and improved blood pressure control [27,28]. For the safety profile, our analysis reveals predominantly mild-to-moderate gastrointestinal effects across all studies.

GLP-RAs mediated delayed gastric emptying and central satiety signaling. The dose-dependent nature of these effects aligns with pharmacodynamic activation of GLP-1 pathways and supports the overall biological plausibility of the observed safety profile.

Our study had some notable strengths: to our knowledge, this is the first and most comprehensive systematic review and meta-analysis of three RCTs on pemvidutide for MASH patients; we included only RCTs to ensure high-quality evidence and enhance the global generalizability of the findings across diverse geographic and ethnic populations.

Also, a subgroup analysis was performed on doses to show the pooled effect of each dose and to clarify the effect of the dose-response relationship of the MASH patients. The consistency in treatment effects across all trials and doses, particularly regarding LFC, ELF, and weight loss, supports the effectiveness of pemvidutide for MASH. We evaluated many outcomes from our pooled trials, including LFC and ELF score as the primary outcome and blood pressure, lipid profile, and safety profile as secondary outcomes.

However, our study had several limitations: 1) There was a clinical heterogeneity among pooled trials, it may be because the difference in follow-up durations 12,24 weeks, and also variability in baseline patient characteristic, particularly regarding background diabetes status requires further stratification in future trials to fully understand the differential efficacy between diabetic and non-diabetic cohorts, although initial data suggests efficacy is preserved regardless of glycemic status. 2) The Phase 1b/2a and Extension trials allowed inclusion of patients with lower fibrosis stages or simple steatosis (NAFLD/MASLD), whereas the IMPACT trial strictly enrolled patients with biopsy-confirmed MASH and fibrosis stages F2-F3. Comparing biomarker changes (like ALT or cT1) across these populations introduces a confounding variable. 3) Two of our included RCTs had a 12-month follow-up, which restricts our ability to draw definitive conclusions regarding long-term fibrosis outcomes compared to 52-week studies.4) Our meta-analysis had only three RCTs, which reduces the statistical power to reliably detect potential publication bias. 5) The limited number of RCTs restricted us from performing any advanced analysis. Future trials with longer follow-up and broader inclusion criteria are needed to evaluate.

## 5. Conclusion

This systematic review and meta-analysis demonstrate that pemvidutide produces substantial reductions in liver fat content and meaningful improvements in cardiometabolic risk factors in patients with obesity and liver disease. Across evaluated dose regimens, pemvidutide consistently reduced hepatic steatosis, body weight, and systolic blood pressure, with additional favorable effects on selected lipid parameters and early signals of fibrosis improvement at lower and intermediate doses.

## Data Availability

All data produced in the present work are contained in the manuscript.

## Supplementary Figures

**Table S1:** Search strategy and results of the databases.

| Databases | Search Strategy | Filters | Results |
| --- | --- | --- | --- |
| **PubMed** | ("pemvidutide" OR "ALT-801" OR "GLP-1 glucagon dual receptor agonist" OR "glucagon-like peptide-1 glucagon dual agonist" OR "GLP-1/glucagon dual agonist")<br>AND<br>("Metabolic Dysfunction-Associated Steatohepatitis" OR "MASH" OR "Non-alcoholic Steatohepatitis" OR "NASH" OR "Non-alcoholic Fatty Liver Disease" OR "Fatty Liver") | All | 121 |
| **Scopus** | ("pemvidutide" OR "ALT-801" OR "GLP-1 glucagon dual receptor agonist" OR "glucagon-like peptide-1 glucagon dual agonist" OR "GLP-1/glucagon dual agonist")<br>AND<br>("Metabolic Dysfunction-Associated Steatohepatitis" OR "MASH" OR "Non-alcoholic Steatohepatitis" OR "NASH" OR "Non-alcoholic Fatty Liver Disease" OR "Fatty Liver") | Title,<br>abstracts,<br>and<br>keywords | 31 |
| **WOS** | ("pemvidutide" OR "ALT-801" OR "GLP-1 glucagon dual receptor agonist" OR "glucagon-like peptide-1 glucagon dual agonist" OR "GLP-1/glucagon dual agonist")<br>AND<br>("Metabolic Dysfunction-Associated Steatohepatitis" OR "MASH" OR "Non-alcoholic Steatohepatitis" OR "NASH" OR "Non-alcoholic Fatty Liver Disease" OR "Fatty Liver") | All | 16 |
| **Cochrane** | ("pemvidutide" OR "ALT-801" OR "GLP-1 glucagon dual receptor agonist" OR "glucagon-like peptide-1 glucagon dual agonist" OR "GLP-1/glucagon dual agonist")<br>AND<br>("Metabolic Dysfunction-Associated Steatohepatitis" OR "MASH" OR "Non-alcoholic Steatohepatitis" OR "NASH" OR "Non-alcoholic Fatty Liver Disease" OR "Fatty Liver") | All | 18 |

**Figure S1.**
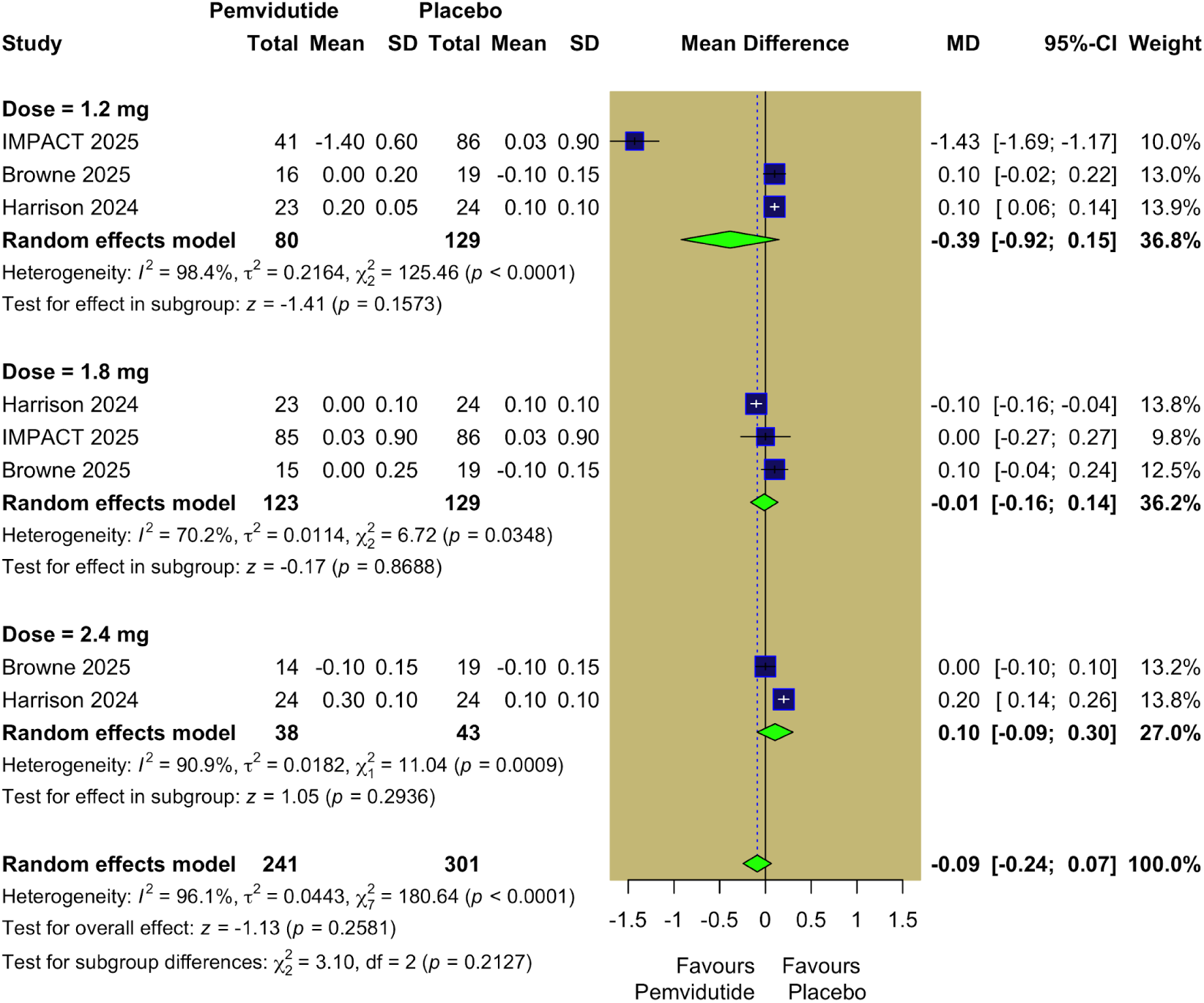
Change in HbA1c (%).

**Figure S2.**
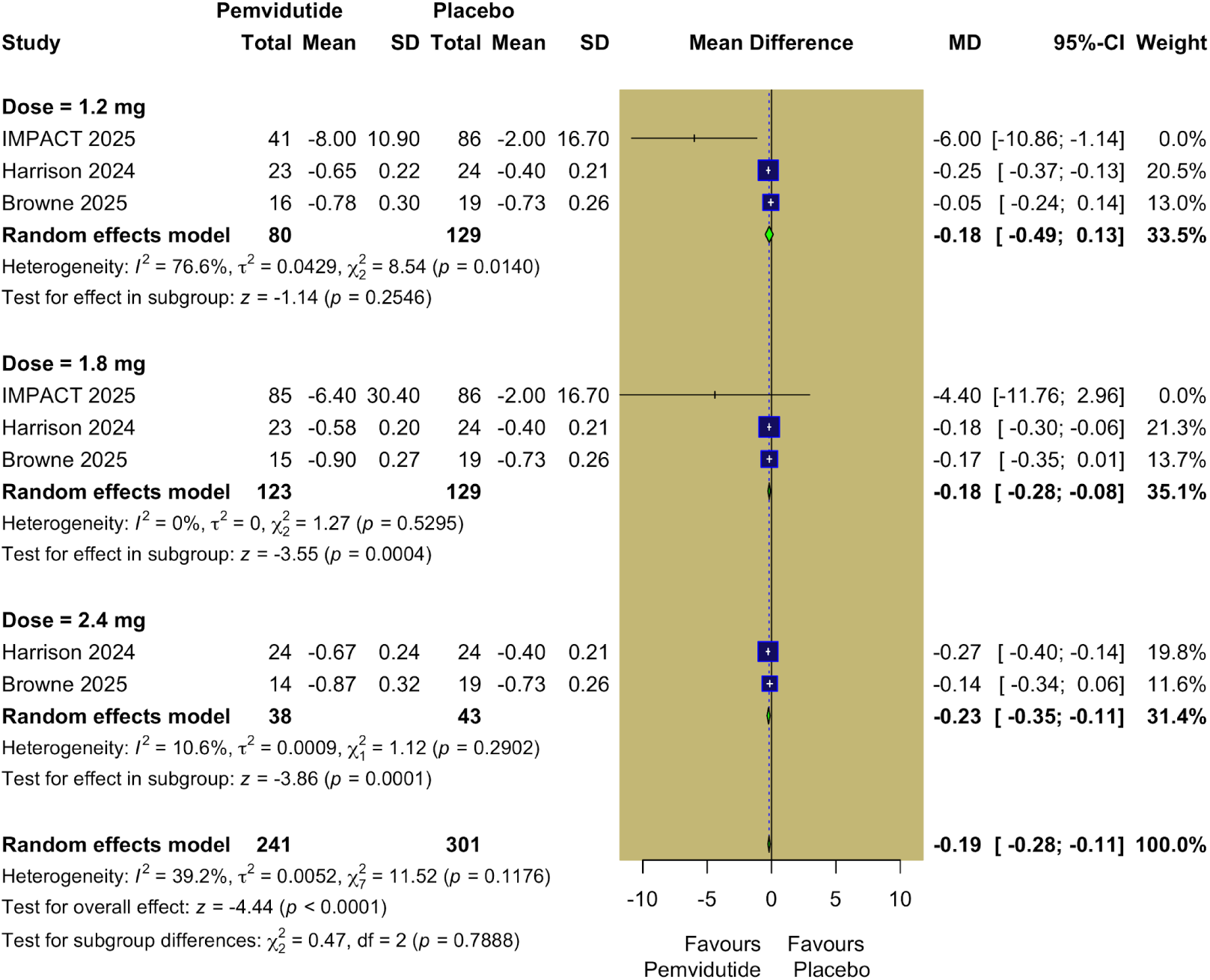
Change in Total Cholesterol (mmol/L).

**Figure S3.**
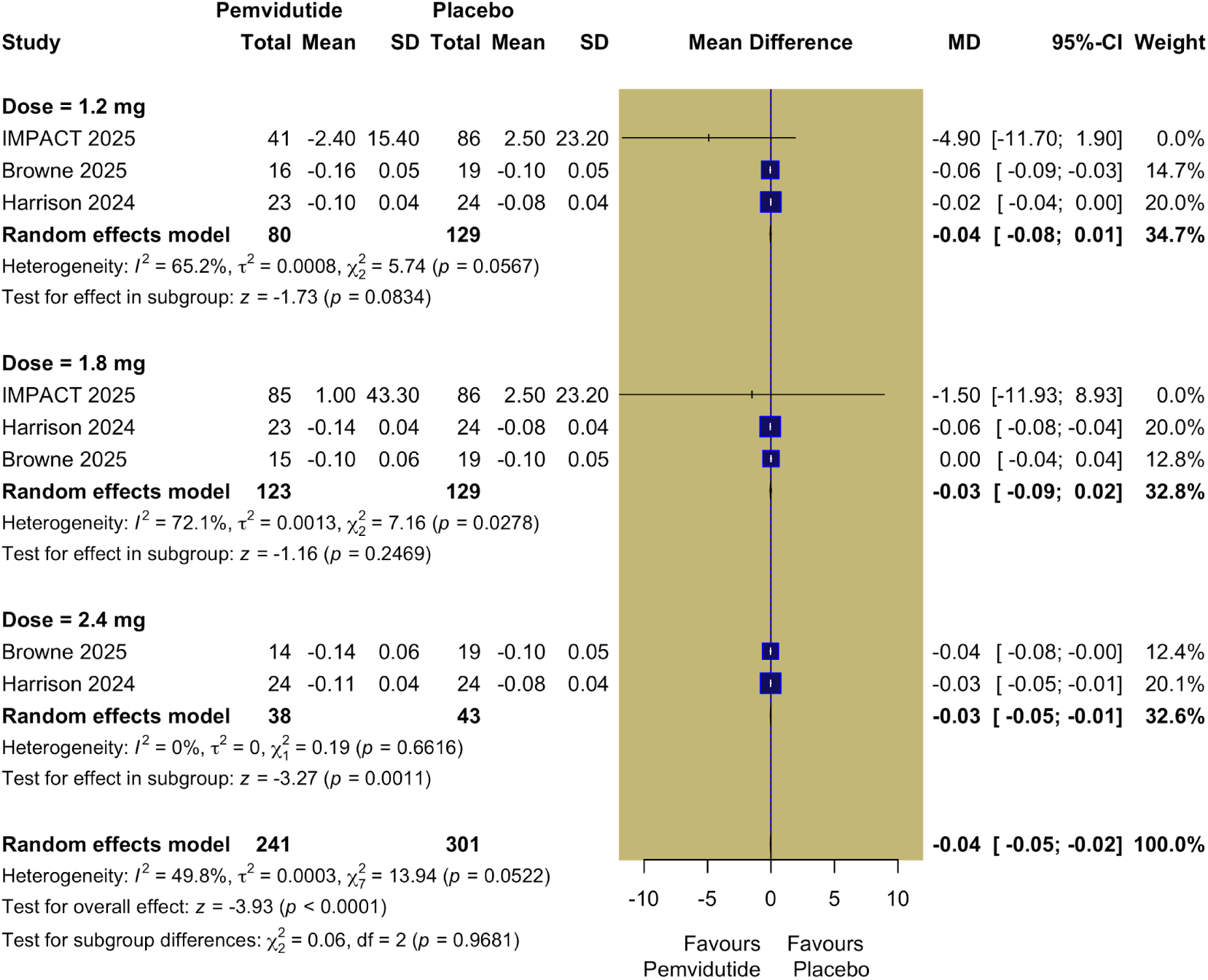
Change in HDL Cholesterol (mmol/L).

**Figure S4.**
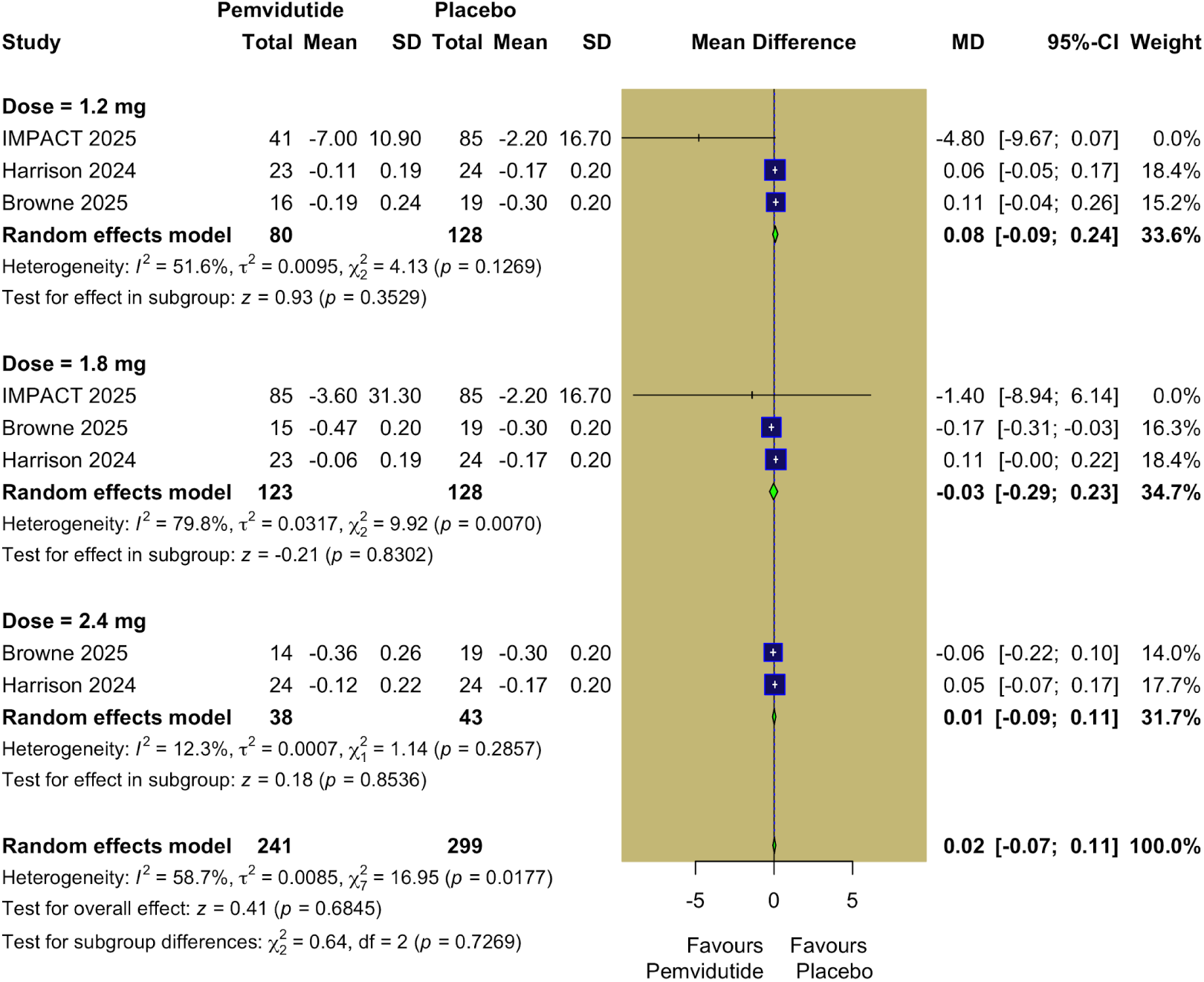
Change in LDL Cholesterol (mmol/L).

**Figure S5.**
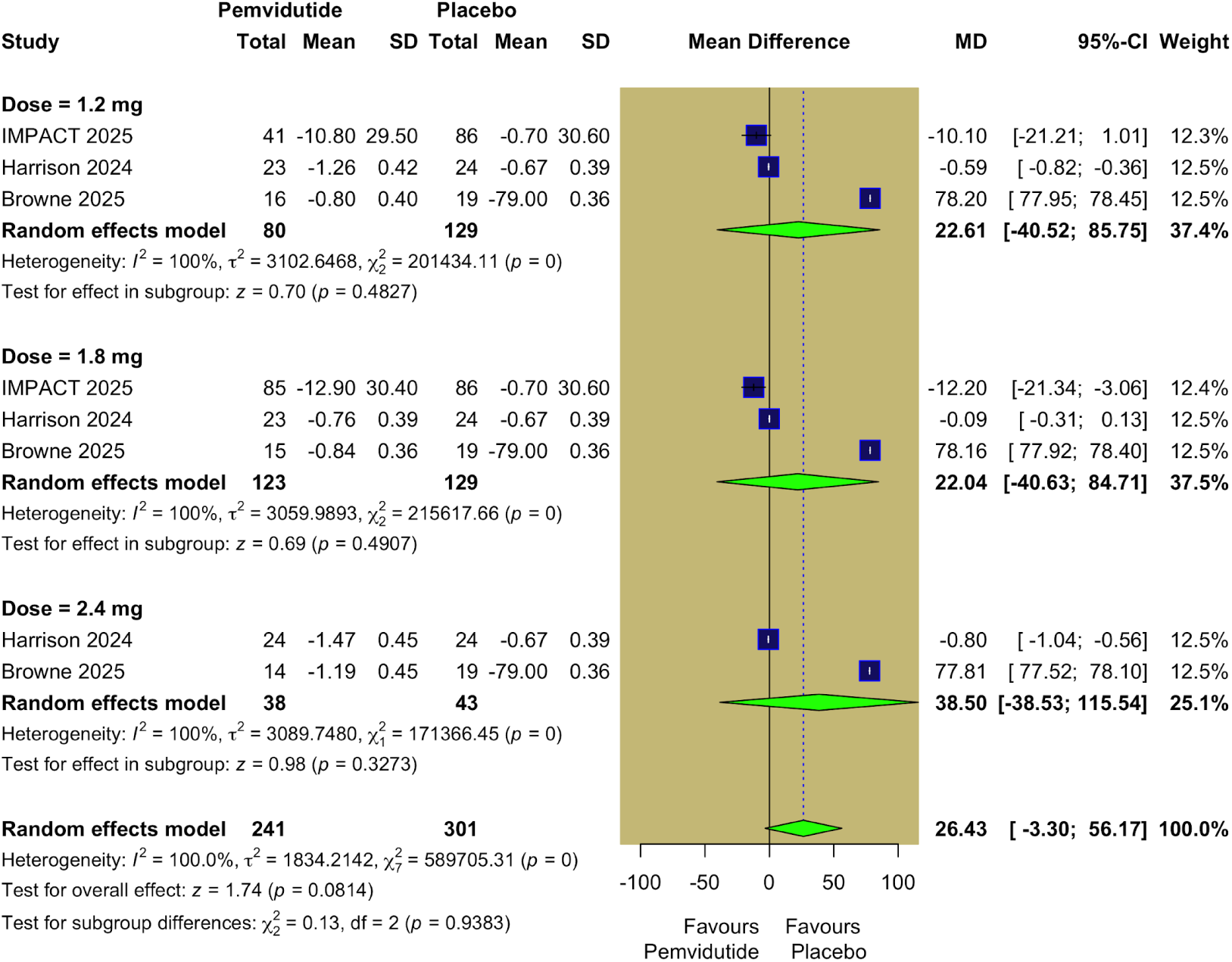
Change in Triglycerides (mmol/L).

**Figure S6.**
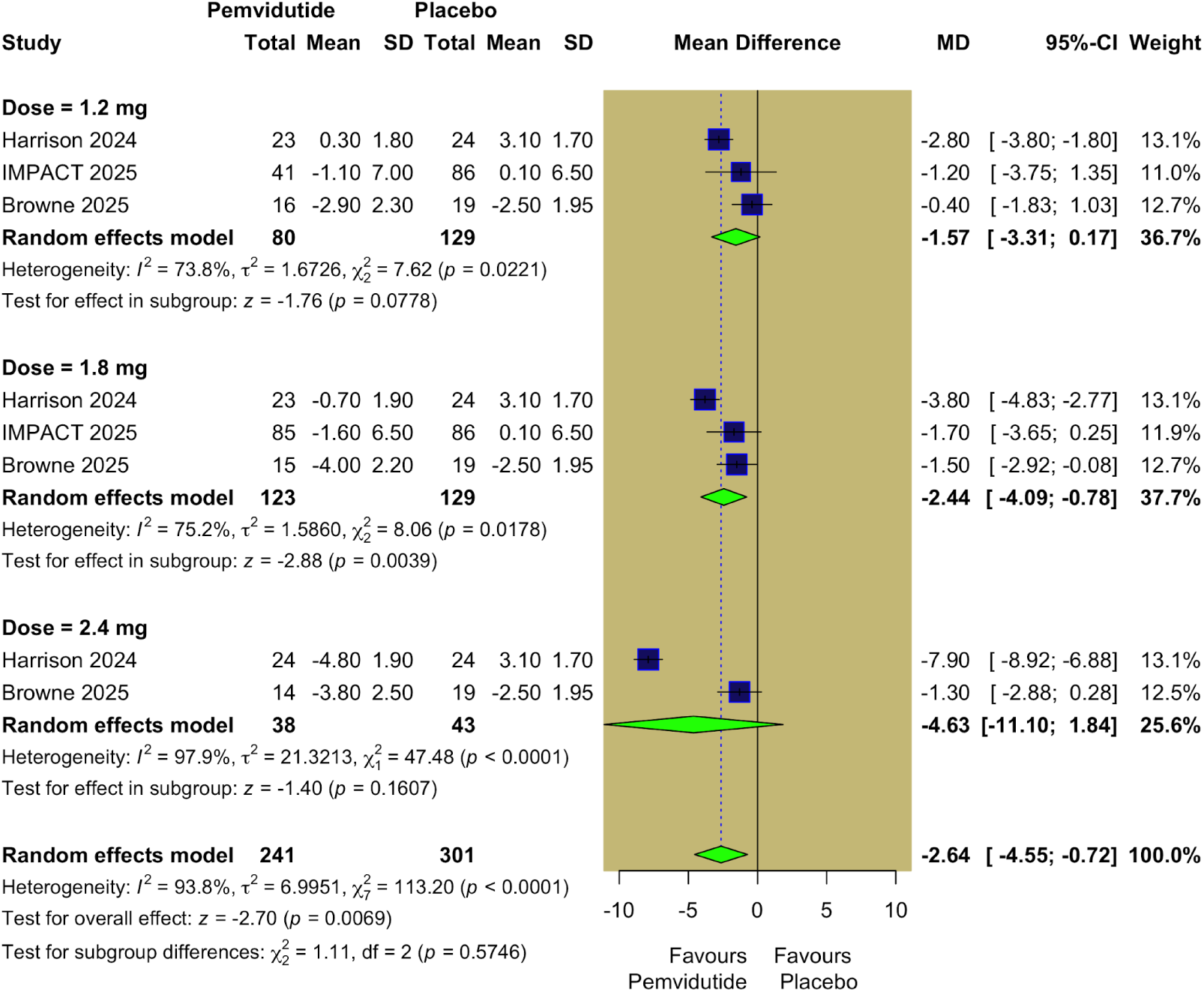
Change in Diastolic Blood Pressure (mm Hg).

**Figure S7.**
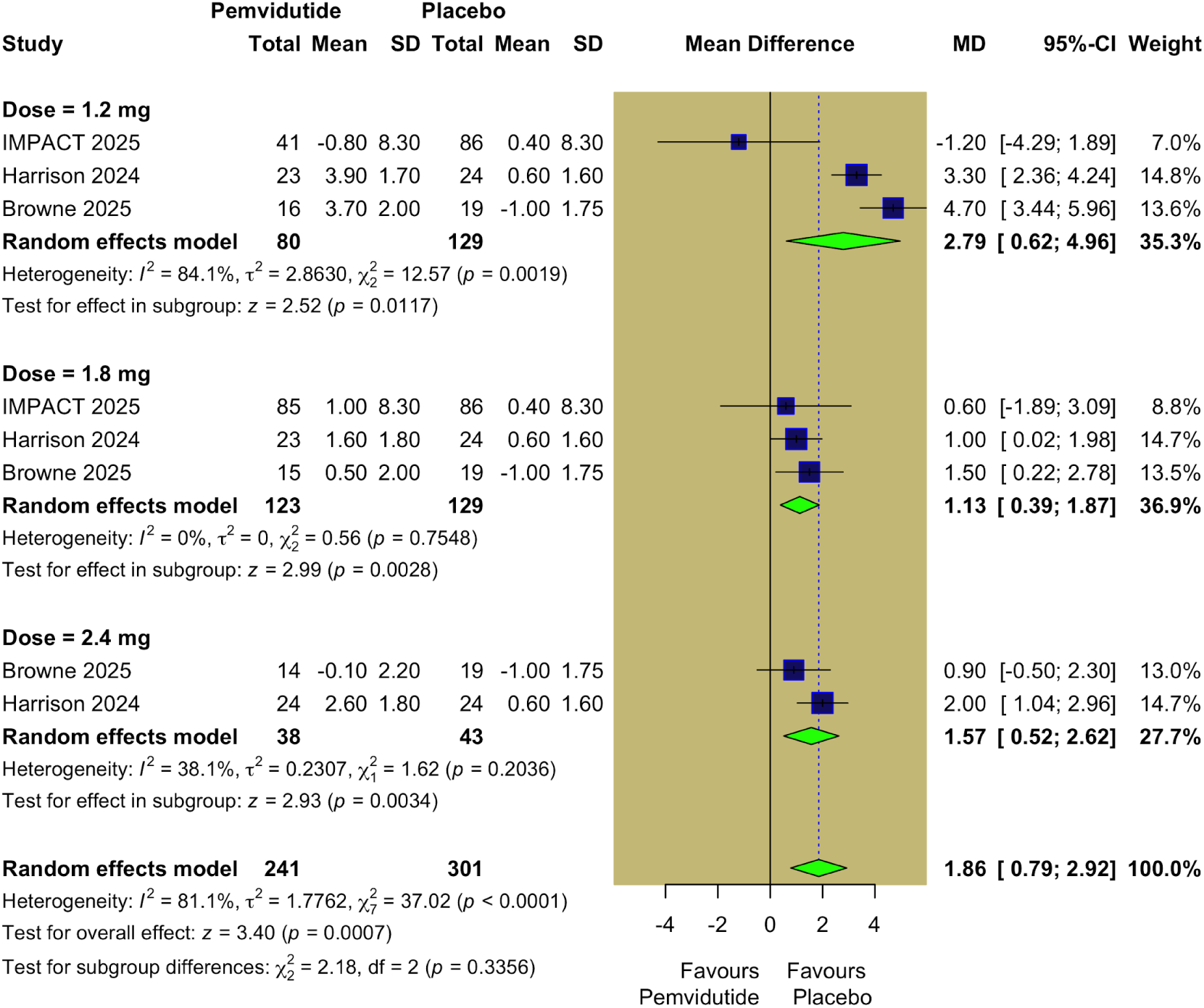
Change in Heart Rate (beats per minute).

**Figure S8.**
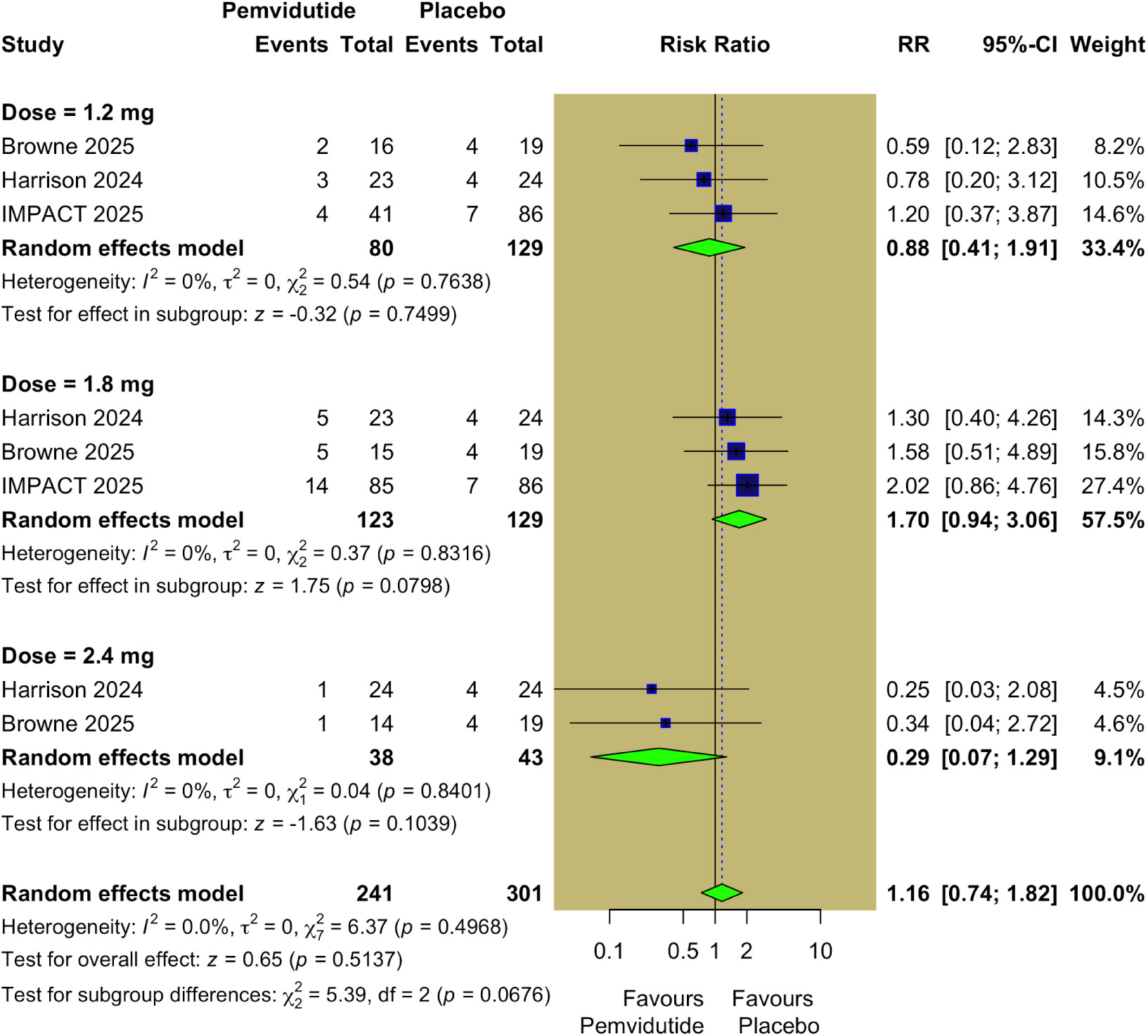
Mild diarrhea.

**Figure S9.**
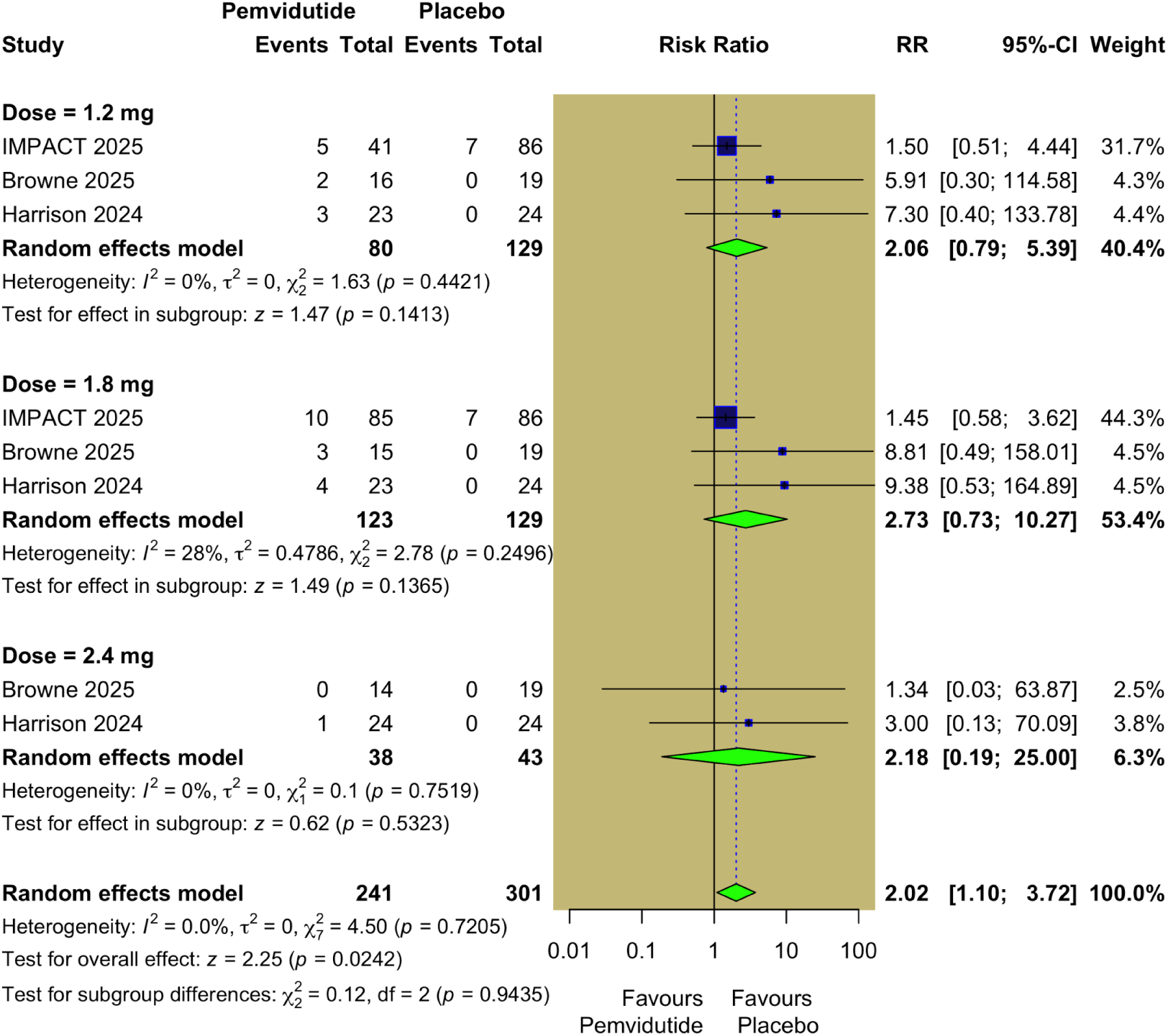
Mild constipation.

**Figure S10.**
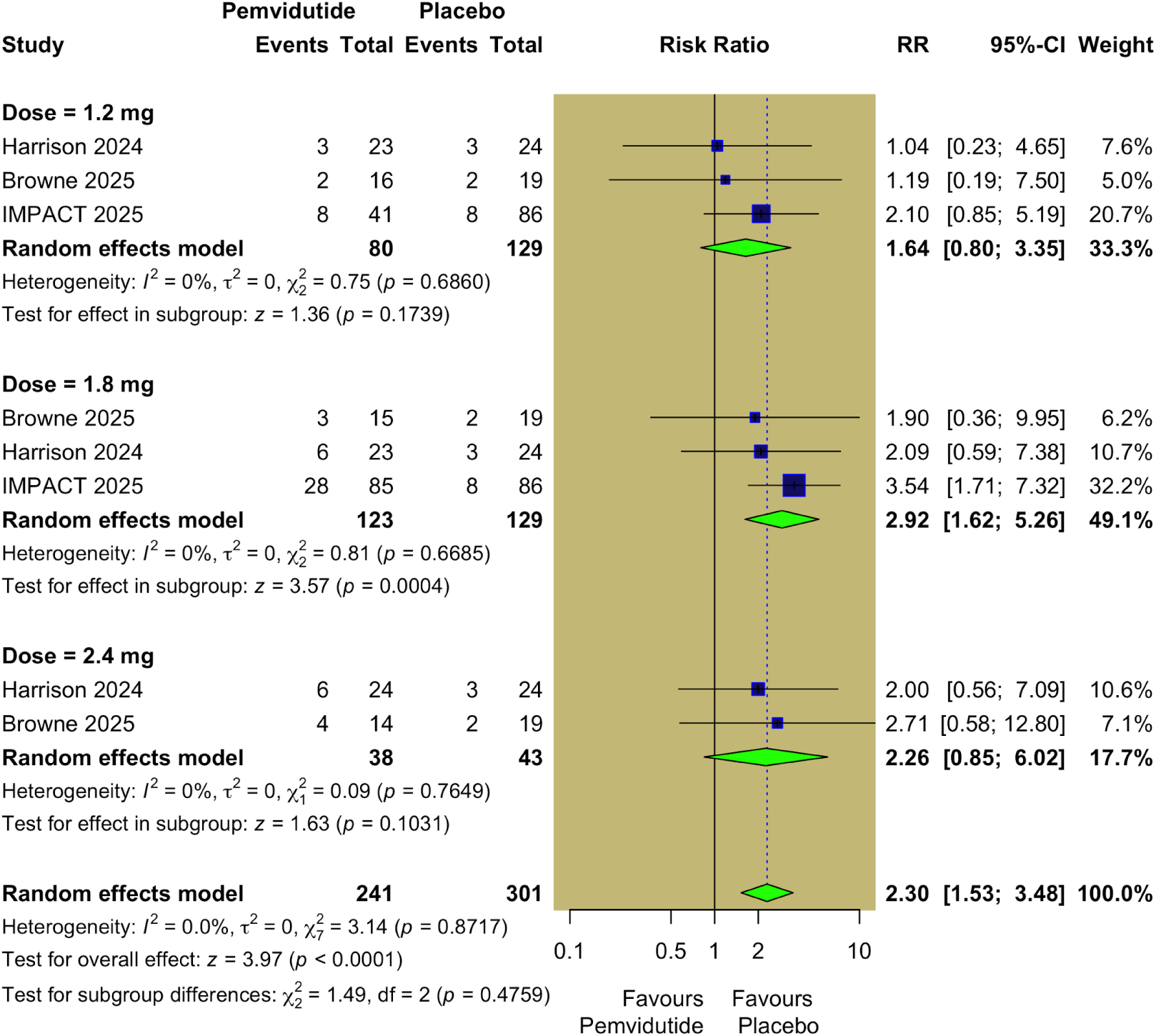
Mild nausea.

**Figure S11.**
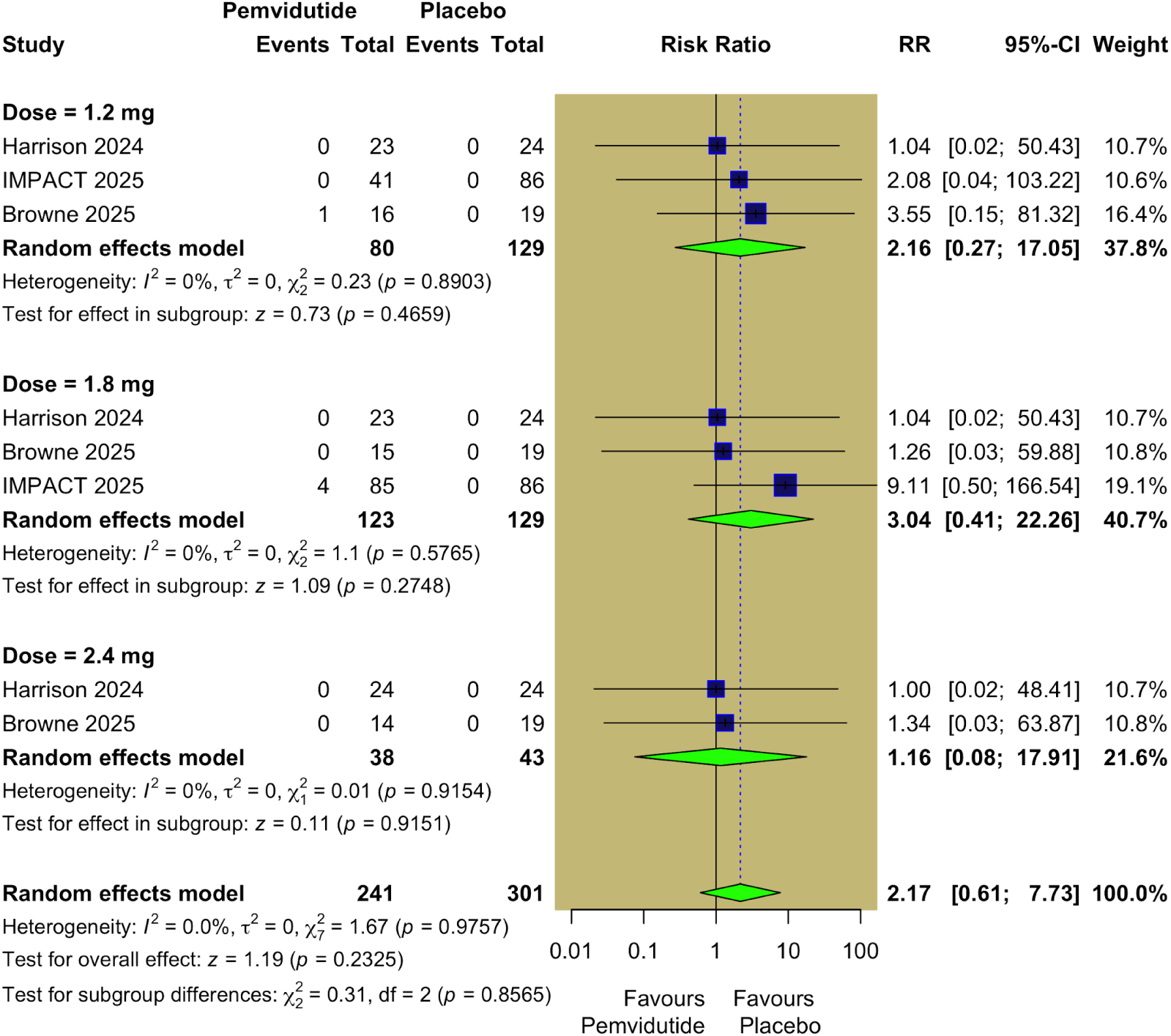
Moderate diarrhea.

**Figure S12.**
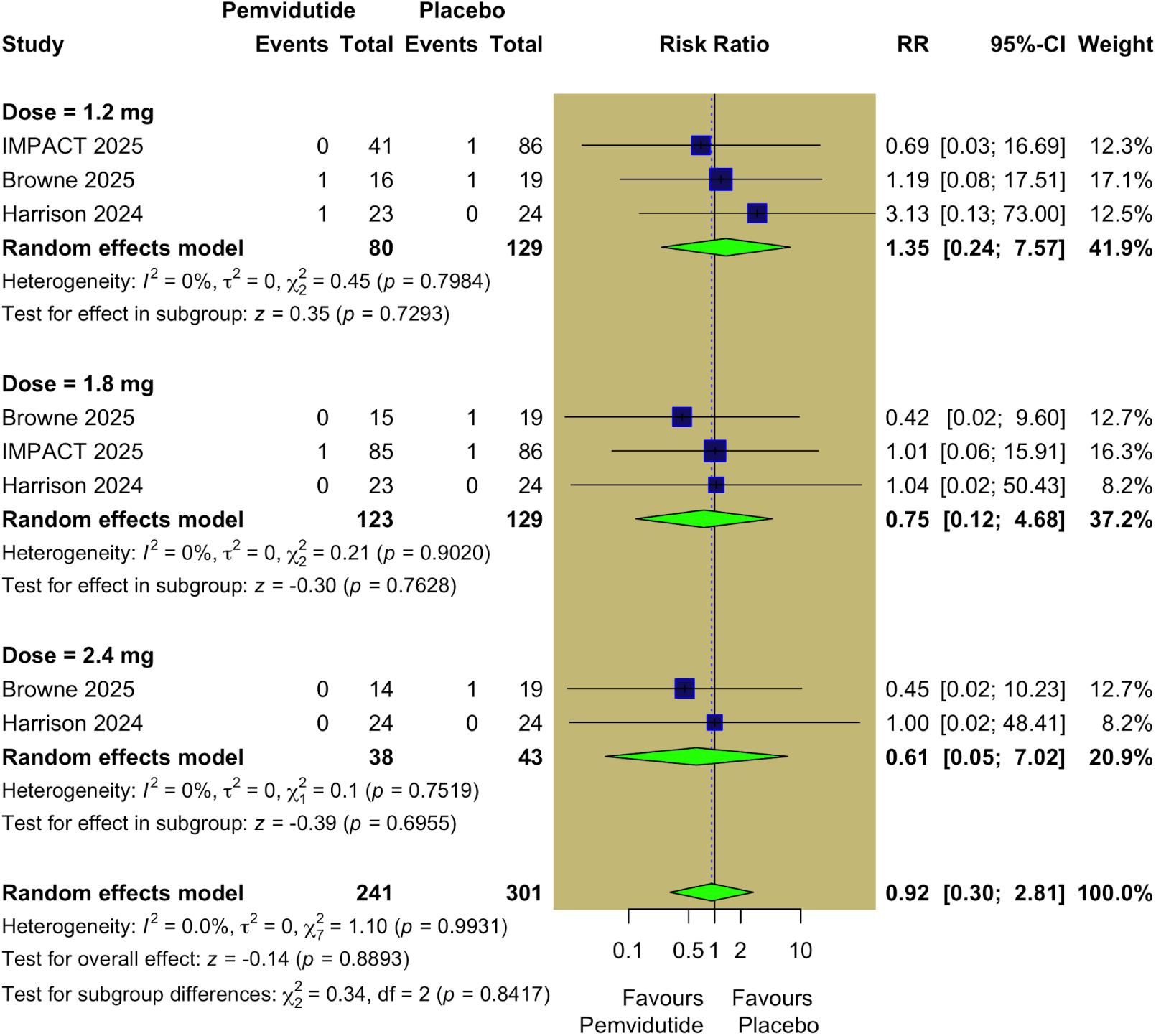
Moderate constipation.

**Figure S13.**
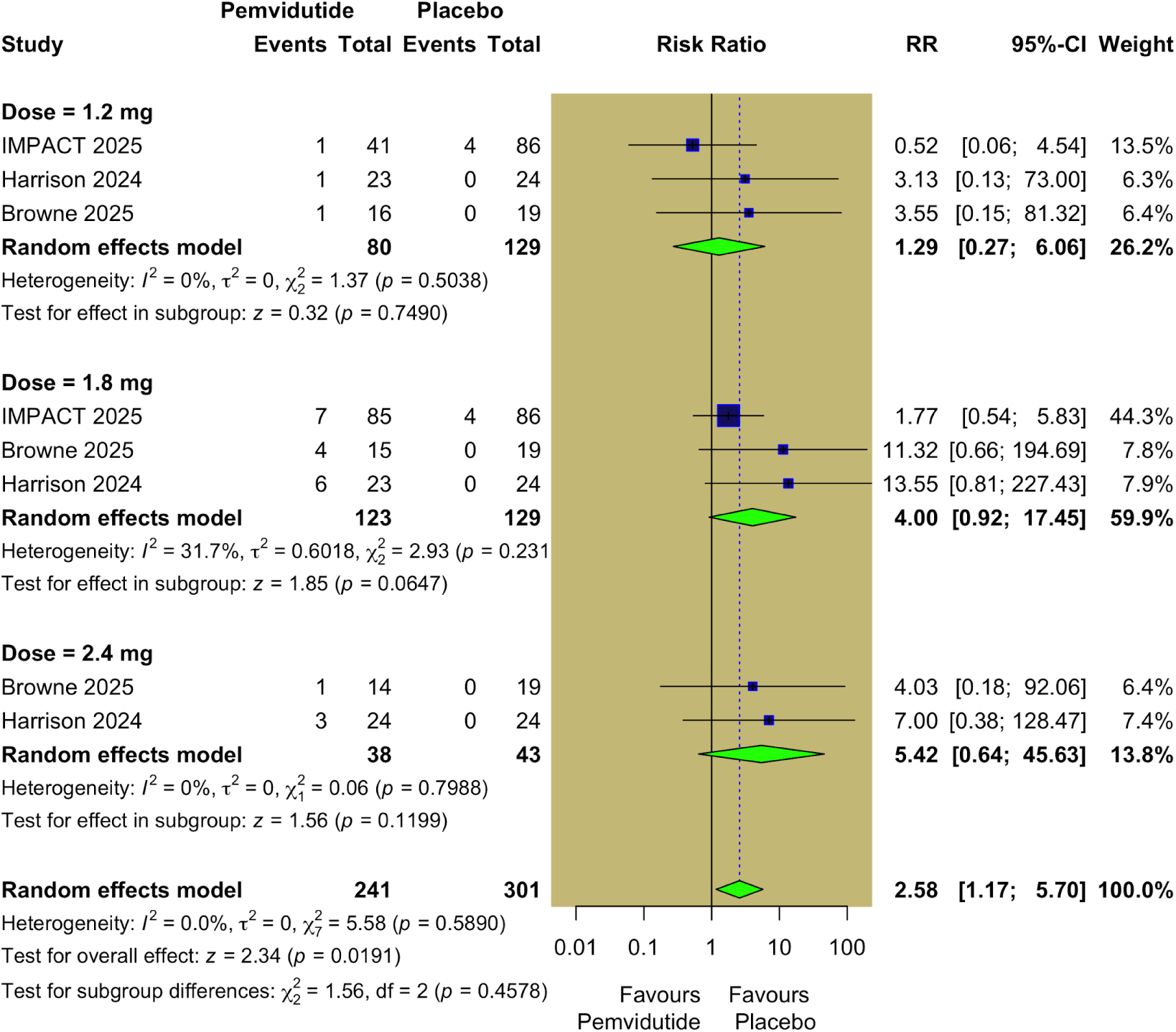
Moderate nausea.

